# Gender gap for accelerometry-based physical activity across different age groups in five Brazilian cohort studies

**DOI:** 10.1101/2023.07.28.23293328

**Authors:** Luiza I. C. Ricardo, Andrea Wendt, Debora Tornquist, Helen Gonçalves, Fernando Wehrmeister, Bruna Gonçalves C. da Silva, Luciana Tovo-Rodrigues, Iná Santos, Aluisio Barros, Alicia Matijasevich, Pedro C. Hallal, Marlos Domingues, Ulf Ekelund, Renata M. Bielemann, Inácio Crohechemore-Silva

## Abstract

**Objectives:** This study aims to evaluate the gender inequalities in accelerometer-based physical activity (PA) across different age groups using data from five Pelotas (Brazil) cohorts.

**Methods:** The data comes from four birth cohort studies, covering all live births in the urban area of Pelotas for each respective year (1982, 1993, 2004, and 2015), and the ‘*Como vai?*’ cohort study focusing on 60 years and above. Raw accelerometry data were collected on the non-dominant wrist using GENEActive/Actigraph devices and processed with the GGIR package. Overall PA was calculated at ages 1, 2, 4, 6, 11, 15, 18, 23, 30, and 60+ years, while moderate-to-vigorous PA (MVPA) was calculated from six years onwards. Absolute (difference) and relative (ratio) gender inequalities were calculated and intersectionality between gender and wealth was also evaluated.

**Results:** The sample sizes per cohort ranged from 965 to 3462 participants. The mean absolute gender gap was 19.3 minutes (95%CI: 12.7; 25.9), with the widest gap at 18 years (32.9 minutes; 95%CI: 30.1; 35.7) for MVPA. The highest relative inequality was found in older adults (ratio 2.0; 95%CI 1.92 to 2.08). Our intersectionality results showed that the poorest men being the most active group, accumulating around 60 minutes more MVPA per day compared with the wealthiest women at age 18.

**Conclusion:** Men were more physically active than women in all ages evaluated. PA gender inequalities start at an early age and intensifies in transition periods of life. Relative inequalities were marked among older adults.

**What is already known on this topic:** Gender inequalities in physical activity have been reported globally, but most of the evidence is focused in adolescents and young adults. The literature lacks studies on children and older adults.

**What this study adds:** We present gender inequalities in accelerometer-based physical activity across several age groups, from 1 year olds to older adults.

**How this study might affect research, practice or policy:** Ou study provides a comprehensive description of gender inequalities, identifying key age groups for intervention.

## BACKGROUND

Men and women present different patterns of morbidity and mortality[1]. Apart from biological differences, socially constructed gender norms, and historical gender inequalities in access to health resources, have harmful effects on women’s health, including physical activity (PA)[2]. Therefore, gender norms shape behaviors from early childhood, influencing not only on children’s behaviors but the encouragement received for active play or physical activities[3,4], and contributing to the lower levels of PA observed in girls compared to boys[5]. These gender differences in PA increase with age,[6] since girls experience less socio-ecological beneficial factors at the individual, family, school, and environmental levels, exacerbating gender inequalities in adolescence[6,7]. In adulthood, for most women the double burden of having a job and taking on most of the housework, motherhood, and lack of family support also contribute to lower levels of PA[8].

Gender or sex inequalities in PA levels have been reported in different age groups and countries with different economic contexts[9–13]. Globally, a gender difference in the prevalence of meeting PA recommendations of 7.1 percentage points was reported in adolescence[9], and around 8.3 percentage points in adulthood[14]. This inequality ranges from the first year of life[15] to older age[16]. However, despite evidence that sex is a consistent correlate of PA at different ages,[17] most studies investigating gender inequalities in PA levels focused in adolescents[1,11,12] or adults[10,18]. The literature still lacks studies exploring these inequalities in children and older adults.

Moreover, most studies measure PA through self-report[10–13,18], which is susceptible to social-desirability and information bias. This study aims to evaluate the gender inequalities in PA across different age groups using accelerometer data from the Pelotas (Brazil) cohorts, including birth cohorts and the ‘Como vai?’ cohort study addressing older adults.

## METHODS

### Participants

We are using data from five cohort studies conducted in Pelotas (Brazi), a city a population of 343.826 inhabitants in 2021 and a GDP per capita of R$ 27,586.96 (U$ 5,290.15)[19]. The first birth cohort (BC) was established in 1982 and additional cohorts were set up in 1993, 2004 and 2015, following all live births in the urban area of Pelotas for each respective year. These studies aim to monitor and examine associations between lifestyle, environmental and social exposures, and health outcomes across the life course and to examine differences across generations. Furthermore, in 2014 The ‘Como vai?’ study was initially designed as a cross-sectional survey of older adult population living in the urban area of Pelotas, which afterwards became a cohort study. Detailed information on each cohort can be found elsewhere[20–26].

Our study included only follow-ups with information on accelerometry, which resulted in samples with ages: one, two, four (2015 BC); six, 11, 15 (2004 BC); 18, 22 (1993 BC); 30 (1982 BC); and 60 years or more (‘Como vai?’).

### Patient involvement

Not applicable.

### Equity, diversity, and inclusion

Given the populational characteristic of the Pelotas cohort studies, representation by gender, race/ethnicity/culture, socioeconomic level, and marginalized groups is guaranteed by design. Also, our research facilities are accessible to participants’ needs, and our research team provides household interviews and measurements when necessary. Furthermore, our study addresses equity by investigating gender and wealth inequalities.

### Accelerometry

The protocol for the Pelotas cohorts comprises 24 hours of accelerometer data collection for all ages, including sleep and water activities. For one- and two-year-olds the accelerometer was placed on the left wrist, and data was collected for two days[27]. Four-year-old children wore the device on their left wrist for seven days, while older participants wore it on the non-dominant wrist for seven days. In the six-, 18- and 30-year follow-ups of 2004, 1993, and 1982 birth cohorts, participants wore the accelerometer on the non-dominant wrist from four to seven days[28].

In the earlier accelerometry data collections (six, 18, 30, and 60+ years) GENEActive (Activinsights, Kimbolton, UK) was used, while in more recent follow-ups the device was the Actigraph GT3X/GT3X+ (Actigraph, Pensacola, USA). High comparability between these brands has previously been reported when raw data is analyzed[29,30]. For this reason, we analyzed the raw signal, which are less affected by company-specific filters.

Data were processed with the GGIR package (version 2.2) in R software[31]. This process included verification of sensor calibration error, detection of sustained abnormally high values, and non-wear detection[32,33]. Vector magnitude was calculated using Euclidian Norm Minus One (ENMO) metric to summarize acceleration from axes x, y, and z into a single-dimensional signal (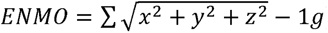). Data was collected and analyzed in a five-second epoch.

To define the minimum number of measurement days included, we used the Spearman-Brown formula, which is based on the Intraclass Correlation Coefficient (ICC) combined with the Spearman-Brown prophecy. Balancing the reliability values and the number of losses, we considered a minimum of one day for one-year-old infants, two days for four-, six-, 18-, and 30-year-old participants, and three days for other ages. This method was described in detail in previous publications[27,34].

The two main outcome variables used in our study were overall PA and moderate to vigorous PA (MVPA). Overall PA was defined as the average Euclidian Norm Minus One (ENMO) per day, expressed in m*g*. MVPA was defined as the average of minutes spent above the threshold of 100m*g* per day[35], using five-minute bouts. We presented both outcome measures to better represent PA behavior in our sample, including the total volume of PA (overall PA) and structured PA (MVPA). The MVPA variable was calculated for individuals aged six years or older.

### Statistical analysis

To illustrate the gender gap in PA, we presented the means of overall PA and MVPA in men and women (sex assigned at birth) for each follow-up in an equiplot (<u>equidade.org/equiplot</u>). The means, 95% confidence intervals (CI) and T-test results are shown in the supplementary material.

Absolute inequalities (gender differences) were calculated by subtracting women’s mean PA (overall PA and MVPA) from men’s mean PA, with positive values indicate a higher mean among men. Relative inequalities were calculated as the ratio between women’s and men’s mean PA, where values above 1 indicate higher PA mean among men. For both absolute and relative inequalities, 95%CIs were calculated using the bootstrap method, which provides the standard errors based on a resampling strategy.

We also analyzed our data stratified for socioeconomic status, aiming to illustrate the PA intersectionality between gender and wealth. The concept of intersectionality, first introduced by Collins (1990)[36], refers to the simultaneous overlapping of multiple forms of inequalities[37]. We used a wealth index using items such as house characteristics and household assets, extracting the first component of a principal component analysis[38]. This component was divided into quintiles, with the first (Q1) representing the poorest group and the last (Q5) representing the wealthiest group. We present means of overall PA and MVPA variables in Q1 and Q5 for men and women in an equiplot. The means, 95%CI, and p-values obtained through t-tests for the intersectionality analysis are available in the supplementary material.

We used a meta-analysis approach with random effect to graphically represent gender inequalities in overall PA and MVPA for each age group. Also, a sensitivity analysis was carried out to verify whether the gender gap is influenced by the amount of PA. To answer this research question, we divided the overall PA (m*g*) and minutes of MVPA in deciles and presented the gender gap in the 10% least active (1^st^ decile) and 10% most active (10^th^ decile). The results of the sensitivity analysis are presented in the supplementary material.

All statistical analyses were performed using Stata 17 (StataCorp. 2021. College Station, TX: StataCorp LLC.)

## RESULTS

The present study includes data from five Pelotas cohort studies, comprising, on average, 2590 participants per follow-up with valid accelerometry data (Table 1). Sample size across cohorts ranged from 965 in the ‘Como vai?’ study to 3462 in the 18-year follow-up (1993BC). Table 1 shows the comparison between the original cohorts and the analytical samples, stratified by gender and wealth quintiles (1^st^ and 5^th^). The 30-year follow-up (1982 BC) had a higher proportion of women (p=0.012) in the analytical sample. None of the remaining comparisons were statistically significant, indicating that the analytical sample is representative of the original cohort when considering gender and wealth distribution.

**Table 1.** Description of original cohorts and analytical samples for all five cohort studies, stratified by gender and wealth quintiles.

| Cohorts' follow-ups | Gender |  |  |  | Wealth quintiles* and gender |  |  |  |  |  |  |  |
| --- | --- | --- | --- | --- | --- | --- | --- | --- | --- | --- | --- | --- |
|  |  |  |  |  | Poorest (Q1) |  |  |  | Wealthiest (Q5) |  |  |  |
|  | All | Men | Women | p-value | All | Men | Women | p-value | All | Men | Women | p-value |
|  | N | N (%) | N (%) |  | N | N (%) | N (%) |  | N | N (%) | N (%) |  |
| 2015 Cohort |  |  |  |  |  |  |  |  |  |  |  |  |
| Original cohort (reference) | 4275 | 2164 (50.6) | 2111 (49.4) | - | 826 | 429 (51.9) | 397 (48.1) | - | 825 | 439 (53.2) | 386 (46.8) | - |
| 1 year | 2974 | 1550 (52.1) | 1424 (47.9) | 0.214 | 590 | 313 (53.1) | 277 (46.9) | 0.706 | 507 | 284 (56.0) | 223 (44.0) | 0.336 |
| 2 years | 2645 | 1363 (51.5) | 1282 (48.5) | 0.473 | 507 | 264 (52.1) | 243 (47.9) | 1.000 | 461 | 253 (54.9) | 208 (45.1) | 0.600 |
| 4 years | 2955 | 1493 (50.5) | 1462 (49.5) | 0.943 | 578 | 305 (52.8) | 273 (47.2) | 0.786 | 523 | 288 (55.1) | 235 (44.9) | 0.537 |
| 2004 Cohort |  |  |  |  |  |  |  |  |  |  |  |  |
| Original cohort (reference) | 4228 | 2193 (51.9) | 2035 (48.1) | - | 707 | 359 (50.8) | 348 (49.2) | - | 653 | 349 (53.4) | 304 (46.6) | - |
| 6 years | 2604 | 1341 (51.5) | 1263 (48.5) | 0.784 | 445 | 227 (51.0) | 218 (49.0) | 0.952 | 402 | 211 (52.5) | 191 (47.5) | 0.799 |
| 11 years | 3348 | 1722 (51.4) | 1626 (48.6) | 0.711 | 553 | 277 (50.1) | 276 (49.9) | 0.821 | 518 | 274 (52.9) | 244 (47.1) | 0.860 |
| 15 years | 1484 | 738 (49.7) | 746 (50.3) | 0.165 | 230 | 115 (50.0) | 115 (50.0) | 0.879 | 246 | 117 (47.6) | 129 (52.4) | 0.117 |
| 1993 Cohort |  |  |  |  |  |  |  |  |  |  |  |  |
| Original cohort (reference) | 5248 | 2603 (49.6) | 2645 (50.4) | - | 863 | 445 (51.6) | 418 (48.4) | - | 856 | 436 (50.9) | 420 (49.1) | - |
| 18 years | 3462 | 1710 (49.4) | 1752 (50.6) | 0.861 | 650 | 339 (52.2) | 311 (47.8) | 0.835 | 640 | 310 (48.4) | 330 (51.6) | 0.347 |
| 23 years | 2783 | 1350 (48.5) | 1433 (51.5) | 0.360 | 517 | 259 (50.1) | 258 (49.9) | 0.617 | 530 | 255 (48.1) | 275 (51.9) | 0.320 |
| 1982 Cohort |  |  |  |  |  |  |  |  |  |  |  |  |
| Original cohort (reference) | 5913 | 3037 (51.4) | 2876 (48.6) | - | 999 | 528 (52.9) | 471 (47.1) | - | 794 | 378 (47.6) | 416 (52.4) | - |
| 30 years | 2680 | 1298 (48.4) | 1382 (51.6) | 0.012 | 489 | 235 (48.1) | 254 (51.9) | 0.087 | 332 | 147 (44.3) | 185 (55.7) | 0.326 |
| COMO VAI? Study |  |  |  |  |  |  |  |  |  |  |  |  |
| Original cohort (reference) | 1451 | 537 (37.0) | 914 (63.0) | - | 282 | 94 (33.3) | 188 (66.7) | - | 273 | 109 (39.9) | 164 (60.1) | - |
| ≥ 60 years | 965 | 364 (37.7) | 601 (62.3) | 0.731 | 180 | 56 (31.1) | 124 (68.9) | 0.684 | 181 | 73 (40.3) | 108 (59.7) | 1.000 |

\*Wealth quintiles at birth for the 2015 and 2004 cohorts, at 2 years for the 1982 cohort, at 11 years for the 1993 cohort, and at baseline for the COMO VAI? study

The gender gap for overall PA and MVPA is illustrated in Figure 1. For overall PA, the gap starts among 1-year-old infants and increasingly widens as PA levels rise, up to 11 years, when the gender gap peaked. There is a drastic decrease in overall PA between 11 and 15 years for both genders, although men still presented higher levels of PA, followed by some stability throughout adulthood. Among older adults, the PA decreases to a lower level than in infants and there is no longer a gender gap (Figure 1 A).

**Figure 1.**
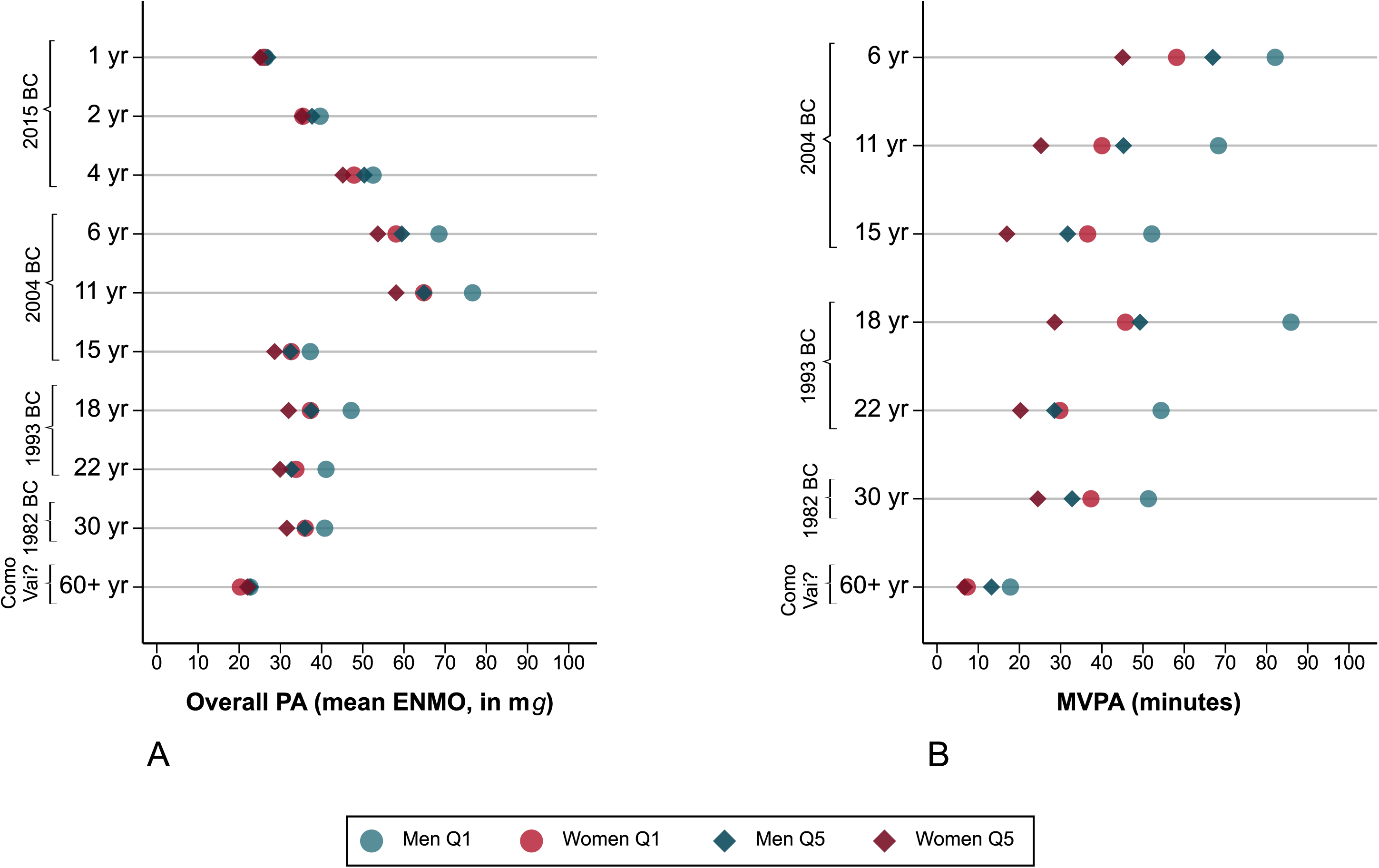
Gender gap for overall PA expressed in m*g* (A) and minutes spent in MVPA (B) among participants of five Pelotas cohort studies.

Regarding MVPA (Figure 1 B), from 6 to 15 years there is a declining pattern for PA with stability in the gender gap. At the age of 18, adolescent men showed an increase in MVPA, which is not followed by women, resulting in the widest gender gap. Between the ages of 22 and 30 years, both MVPA and gender gap show little variation, and a steep decrease is observed for older adults, although the gender gap remains, with older men spending more time in MVPA than older women.

The absolute gender inequalities for overall PA and MVPA are described in Figure 2. For overall PA (Figure 1 A), the widest gender gap was found at 11 years (Difference 9.8 m*g*; 95% CI 8.7 to 11.0), while for MVPA (Figure 1 B) the widest difference was found at 18 years (Difference 32.9 minutes; 95% CI 30.1 to 35.7). For both overall PA and MVPA, men had higher PA than women in virtually all ages, although there was no gap between men and women in overall PA among older adults (Figure 2 A).

**Figure 2.**
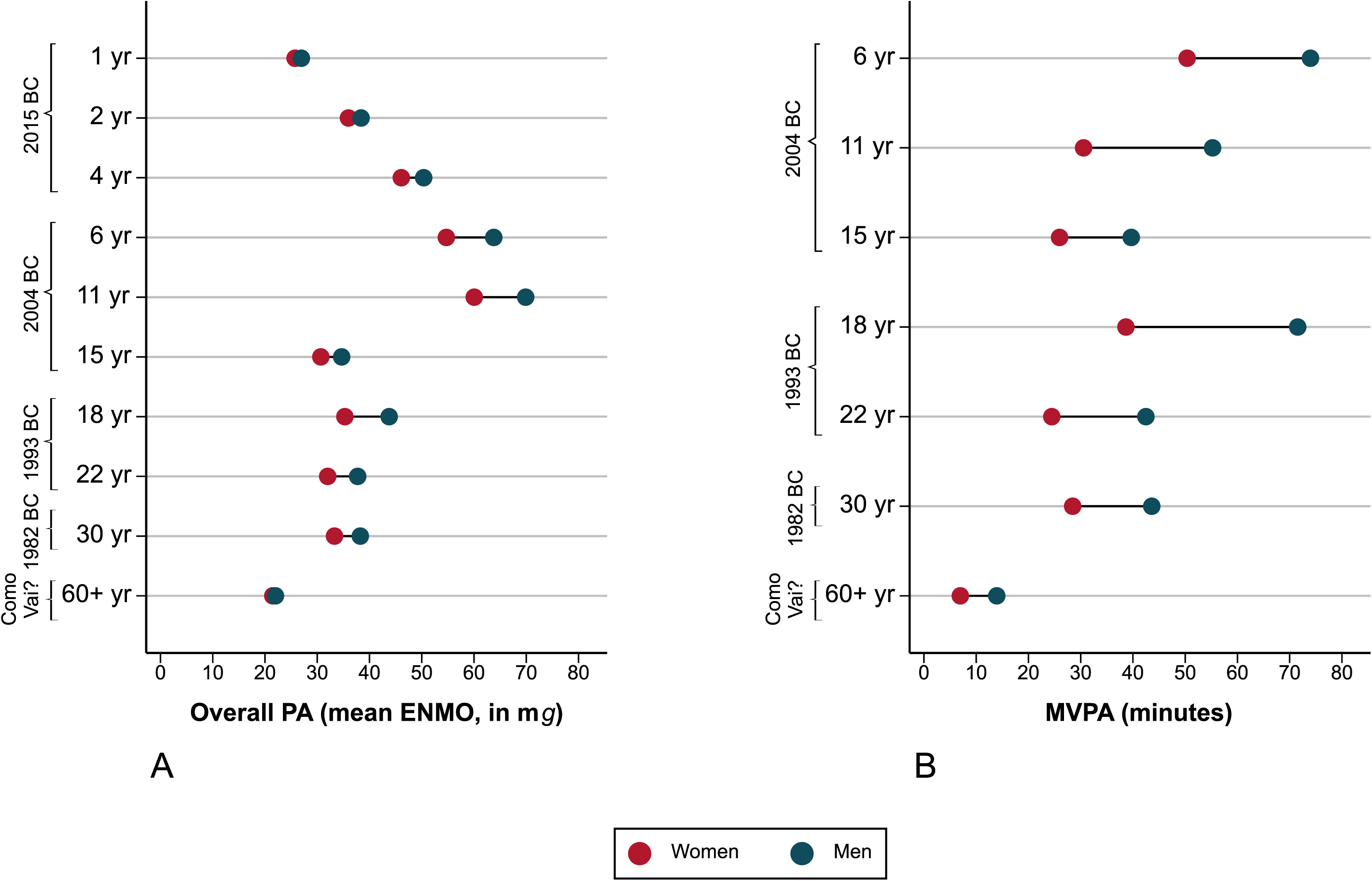
Absolute gender inequalities for overall PA expressed in m*g* (A) and minutes spent in MVPA (B) among participants of five Pelotas cohort studies.

Regarding relative gender inequalities (Figure 3 A and B), 18-year-olds presented the highest gender inequalities for overall PA (ratio 1.24; 95%CI 1.22 to 1.26), while older adults presented the lowest inequalities (ratio 1.02; 95%CI 0.98 to 1.07) although not statistically significant. On the other hand, for MVPA older adults presented the highest relative inequalities (ratio 2.0; 95%CI 1.92 to 2.08), while 6-year-olds had the lowest inequalities (ratio 1.47; 95%CI 1.40 to 1.54).

**Figure 3.**
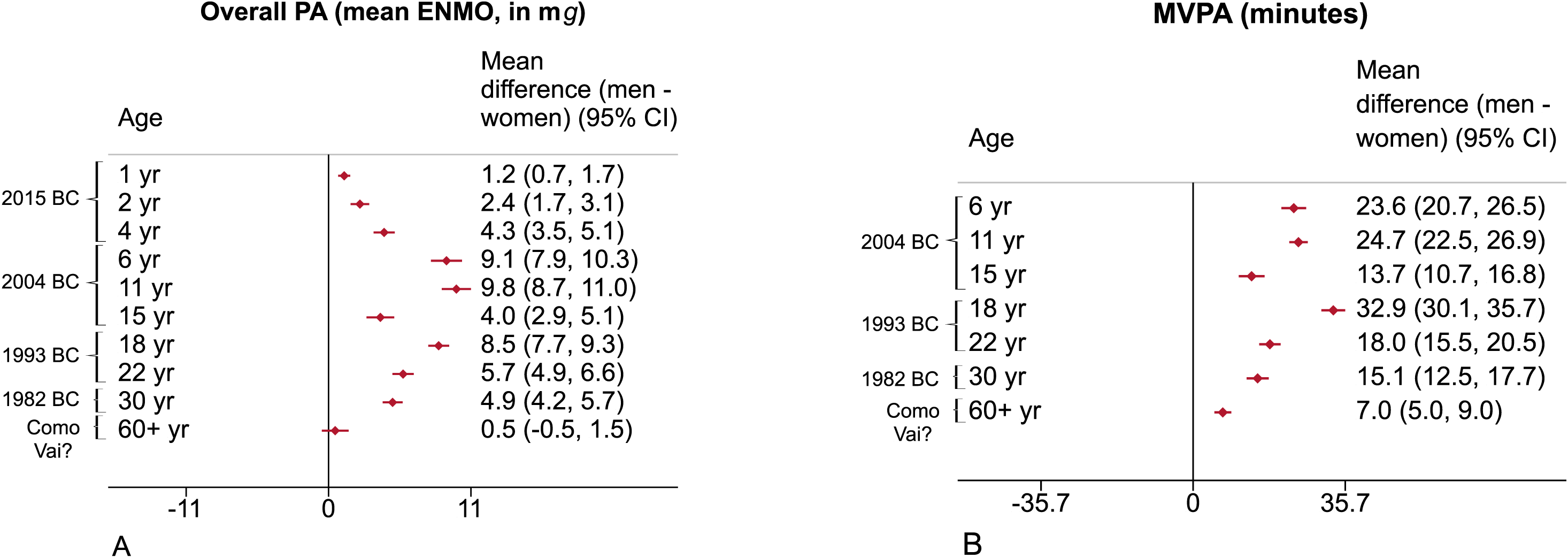
Relative gender inequalities for overall PA expressed in m*g* (A) and minutes spent in MVPA (B) among participants of five Pelotas cohort studies.

Figure 4 shows the gender gap for PA across the first (Q1, 20% poorest) and last (Q5, 20% richest) wealth quintiles. The double stratification for overall PA showed the higher acceleration among the poorest groups in all ages, although the wealthiest men have similar overall PA levels to the poorest women from the age of 11 years onwards. The highest gaps are shown at ages 11 and 18 years, with the poorest men showing around 20 m*g* of difference from the wealthiest women (Figure 4 A). For MVPA, during childhood and early adolescence (1 and 11 years) boys showed more MVPA than girls, regardless of wealth. From 15 years to adulthood, the poorest women show similar results to the wealthiest men. The highest gap is at age 18 years when the poorest men show around 60 minutes of MVPA more than the wealthiest women.

**Figure 4.**
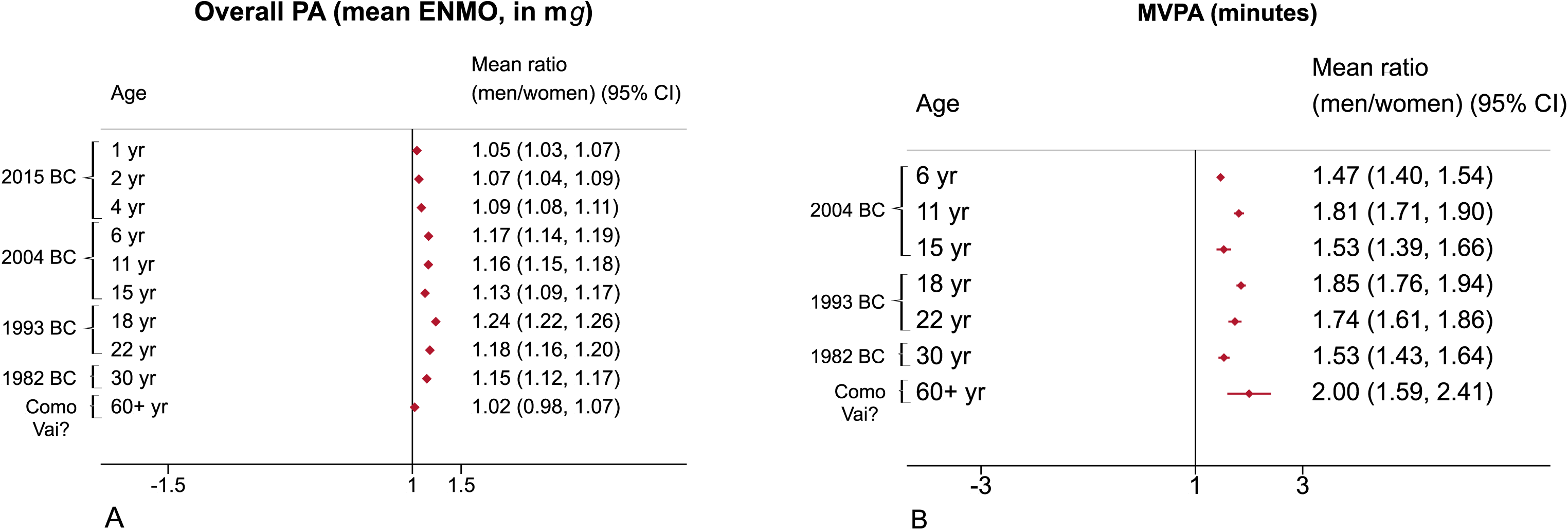
Gender gap for overall PA expressed in m*g* (A) and minutes spent in MVPA (B) between the first and last wealth quintiles among participants of five Pelotas cohort studies.

Gender inequalities in overall PA and MVPA for the highest and lowest PA deciles are shown in Supplementary Figure 1. The highest decile of overall PA (D10) showed increasing gender inequalities from 1 to 11 years, followed by stability during adulthood and a drastic decline among older adults. The widest gap for overall PA in D10 was observed at age 11, with approximately 20 m*g* of difference between boys and girls. The MVPA gap in D10 also increased with age until the end of adolescence (18 years), when the highest gap was found (~70 minutes) and narrowed thereafter.

## DISCUSSION

This work describes the gender gap for PA in five Pelotas cohort studies, including a broad age range of participants. We attempted to describe the patterns of absolute and relative PA gender inequalities in multiple age groups by exploring overall PA and MVPA, aiming at representing both everyday activities and structured PA, such as sports and exercises. Our findings showed that both overall PA and MVPA were consistently higher among men.

We presented absolute and relative measures of inequalities. While absolute measures have easier interpretation, relative measures can better express proportional differences, allowing comparisons at different scales. They are complementary because their interpretability differ according to the magnitude of outcomes occurrence^49^. For example, when there is a lower occurrence there could be marked relative inequalities, but modest absolute inequalities. In our case, older adults presented a low level of MVPA so the absolute inequality measure identified a difference of only 7 minutes between men and women. On the other hand, the relative inequality measure identified a mean of MVPA in men 2 times higher than in women for this group - the highest relative inequality among all age groups. This does not mean that one measure is better than the other, but prefereably, both measures should be shown to improve interpretability and better inform policymakers’ decisions[39].

It is important to underline that overall PA measured by accelerometry reflects any activity performed throughout the day, including daily routine activities and intermittent and sporadic movements, and not just structured PA. Also, PA described in m*g* presents even less interpretability of the data, hampering its translation into daily activities. However, recent evidence has shown that, among inactive adults, the minimum clinically important difference of approximately 1 m*g* corresponds to about 5 to 6 min of brisk walking per day, and an increase in average daily acceleration of this magnitude is associated with reduced risk of all-cause mortality (Hazard Ratio: 0.95; 95%CI: 0.94; 0.96)[40]. When applying this example to our results, the mean absolute gender gap of 5 m*g* found in the present study, would correspond to approximately 25 to 30 minutes of daily brisk walking.

The PA gender gap starts early in life, with inequalities already found in one-year-old infants. The intrinsic societal restrictive gender norms are the main drivers of this discrepancy in PA, which indirectly shapes how parents raise their children, including how to dress a young girl to a play-day, which toys are gifted to boys and girls, and above all, the level of incentive to active play they receive[41]. As a result, young girls often receive less favorable influences at the individual, family, school, and environmental levels[42]. When discussing PA promotion at early ages it is imperative to highlight the tracking effect of PA, where behaviours acquired during childhood are more likely to be carried out to adolescence and adulthood[43].

Another relevant finding is that transition phases, from childhood to adolescence (11 years old) and from adolescence to early adulthood (18 years old), showed the widest gender inequalities for overall PA and MVPA, respectively. Some of the barriers surrounding the women’s PA might be more evident in adolescence, especially body dissatisfaction and feeling uncomfortable during the practice, in addition to less social support from peers, family, and teachers than their male counterparts, and less access to safe and welcoming facilities, equipment, transport, and training[44,45]. These factors shape female attitudes towards PA throughout adolescence and could affect this behavior throughout life[44] and this might be intensified by transition life events, such as entering university and/or the job market, and above all motherhood, which often lead women to accumulate even more barriers to PA practice[46,47].

Our results on the PA’s intersectionality between gender and wealth showed higher acceleration among the poorest groups, with the 20% richest women being the least active group, and the 20% poorest men being the most active group. To better interpret this result its imperative to understand that accelerometry-based overall PA includes all PA domains: leisure time, occupational, commuting, and domestic. This is a limitation of accelerometry, since this measure lacks contextual information on the domains of PA, and then we observe controversial wealth patterns for this behavior[48]. In this sense, the literature consistently reports a higher level of domestic, occupational, and commuting PA among socioeconomically vulnerable populations when considering self-reported measures[49]. On the other hand, the richest groups usually have a privileged position in PA practice, being able to pay for private and expensive activities, and taking place during their leisure time[50]. The sensitivity analysis revealed interesting results when stratifying PA levels by gender and the amount of PA. For both outcomes (overall PA and MVPA), we identified a clear pattern of a higher gender gap when looking at the most active group. These results highlight that women, in general, present a much lower level of PA than men, even among the most active group. In other words, the highest decile of PA is a very selected part of the sample (with an average of close to 100 minutes of MVPA). However, we identified a huge difference between most active men and women already at younger ages. Even the most active women are not close to most active men. These findings suggest that the gender gap is also evident in women with greater opportunities, support, and facilitators for PA practice.

### Limitations

Because we described data from five different cohort studies, this analysis is not completely longitudinal. PA levels are from different generations, consequently, conclusions should be based on descriptive assumptions of a wide age range instead of a lifecycle analysis. Also, although raw data minimizes the differences between the two device brands used, some residual distinctions could remain. Howerver, since men and women wore the same device, gender inequalities were not affected. Finally, at 15 years, the follow-up was interrupted by the COVID-19 pandemic, which could have impacted PA results.

### Policy implications

Our findings demonstrated that PA gender inequalities start at an early age and intensify in transition periods of life, but also, relative gender inequalities are marked among older adults. Actions to promote PA in women must consider a life cycle strategy, starting at an early age, intensifying actions at transition periods, but still being mindful of older adults’ needs.

## Contributorship

LICR, AW, and DT drafted substantial parts of the manuscript. LICR also performed data analysis and interpretation. HG, FW, BGCS, LT, IS, AB, AM, PCH, MD, UE, RB, IC served as scientific advisors. All authors critically reviewed and approved the final version of this manuscript.

## Supporting information

Suppelementary material

Unindentified data

## Data Availability

All data relevant to the study are included in the article or uploaded as supplementary information.

https://osf.io/du96j/

## Acknowledgements

The authors would like to thank all members from the *Grupo de Estudos e Pesquisa em Acelerometria* (GEPEA) for the valuable scientific discussions during the conception of this manuscript.

## Ethical approval

The 1982, 1993, and 2004 Pelotas birth cohorts study protocols were approved by the Medical School Ethics Committee of the Federal University of Pelotas (respective registration numbers: 16/12; 1.250.366; 40601116) and the 2015 birth cohort, by the School of Physical Education Ethics Committee of the Federal University of Pelotas (registration 26746414.5.0000.5313). The ‘Como vai?’ study protocol was approved by the Research Ethics Committee of the Faculty of Sciences of the Federal University of Pelotas (registration 24538513.1.0000.5317). The participation of the individuals in each study was voluntary, and informed consent was obtained from all participants.

## REFERENCES

1 de Looze M, Elgar FJ, Currie C, et al. Gender Inequality and Sex Differences in Physical Fighting, Physical Activity, and Injury Among Adolescents Across 36 Countries. Journal of Adolescent Health 2019;64:657–63. doi:10.1016/j.jadohealth.2018.11.007

2 Hawkes S, Buse K. Gender and global health: Evidence, policy, and inconvenient truths. The Lancet 2013;381:1783–7. doi:10.1016/S0140-6736(13)60253-6

3 Nilsen AKO, Anderssen SA, Ylvisaaker E, et al. Physical activity among Norwegian preschoolers varies by sex, age, and season. Scand J Med Sci Sports 2019;29:862–73.

4 Mattocks C, Hines M, Ness A, et al. Associations between sex-typed behaviour at age 3½ and levels and patterns of physical activity at age 12: the Avon Longitudinal Study of Parents and Children. Arch Dis Child 2010;95:509–12.

5 Arts J, Drotos E, Singh AS, et al. Correlates of Physical Activity in 0- to 5-year-olds: A Systematic Umbrella Review and Consultation of International Researchers. Sports Medicine 2023;53:215–40. doi:10.1007/S40279-022-01761-5/TABLES/6

6 Schlund A, Reimers AK, Bucksch J, et al. Sex/gender considerations in school-based interventions to promote children’s and adolescents’ physical activity: A systematic review. German Journal of Exercise and Sport Research 2021;51:257–68. doi:10.1007/S12662-021-00724-8/TABLES/3

7 Telford RM, Telford RD, Olive LS, et al. Why Are Girls Less Physically Active than Boys? Findings from the LOOK Longitudinal Study. PLoS One 2016;11:e0150041. doi:10.1371/JOURNAL.PONE.0150041

8 Evenson KR, Aytur SA, Borodulin K. Physical Activity Beliefs, Barriers, and Enablers among Postpartum Women. J Womens Health 2009;18:1925. doi:10.1089/JWH.2008.1309

9 Guthold R, Stevens GA, Riley LM, et al. Global trends in insufficient physical activity among adolescents: a pooled analysis of 298 population-based surveys with 1·6 million participants. Lancet Child Adolesc Health 2020;4:23–35.

10 Mielke GI, da Silva ICM, Kolbe-Alexander TL, et al. Shifting the Physical Inactivity Curve Worldwide by Closing the Gender Gap. Sports Medicine 2017 48:2 2017;48:481–9. doi:10.1007/S40279-017-0754-7

11 Brazo-Sayavera J, Aubert S, Barnes JD, et al. Gender differences in physical activity and sedentary behavior: Results from over 200,000 Latin-American children and adolescents. PLoS One 2021;16:e0255353. doi:10.1371/JOURNAL.PONE.0255353

12 Ricardo LIC, Wendt A, Costa C dos S, et al. Gender inequalities in physical activity among adolescents from 64 Global South countries. J Sport Health Sci 2022;11:509. doi:10.1016/J.JSHS.2022.01.007

13 Werneck AO, Baldew SS, Miranda JJ, et al. Physical activity and sedentary behavior patterns and sociodemographic correlates in 116,982 adults from six South American countries: The South American physical activity and sedentary behavior network (SAPASEN). International Journal of Behavioral Nutrition and Physical Activity 2019;16:1–11. doi:10.1186/S12966-019-0839-9/FIGURES/3

14 Guthold R, Stevens GA, Riley LM, et al. Worldwide trends in insufficient physical activity from 2001 to 2016: a pooled analysis of 358 population-based surveys with 1·9 million participants. Lancet Glob Health 2018;6:e1077–86.

15 Ricardo LIC, Da Silva ICM, De Andrade Leão OA, et al. Objectively measured physical activity in one-year-old children from a Brazilian cohort: Levels, patterns and determinants. International Journal of Behavioral Nutrition and Physical Activity 2019;16:1–13. doi:10.1186/s12966-019-0895-1

16 Lin YC, Yeh MC, Chen YM, et al. Physical activity status and gender differences in community-dwelling older adults with chronic diseases. J Nurs Res 2010;18:88–97. doi:10.1097/JNR.0B013E3181DDA6D8

17 Aleksovska K, Puggina A, Giraldi L, et al. Biological determinants of physical activity across the life course: a “Determinants of Diet and Physical Activity” (DEDIPAC) umbrella systematic literature review. Sports Medicine - Open 2019 5:1 2019;5:1–18.

18 Wendt A, Ricardo LIC, Costa CS, et al. Socioeconomic and Gender Inequalities in Leisure-Time Physical Activity and Access to Public Policies in Brazil From 2013 to 2019. J Phys Act Health 2021;18:1503–10. doi:10.1123/JPAH.2021-0291

19 IBGE | Cidades@ | Rio Grande do Sul | Pelotas | Panorama. https://cidades.ibge.gov.br/brasil/rs/pelotas/panorama (accessed 20 Sep 2022).

20 Santos IS, Barros AJD, Matijasevich A, et al. Cohort profile: the 2004 Pelotas (Brazil) birth cohort study. Int J Epidemiol 2011;40:1461–8. doi:10.1093/IJE/DYQ130

21 Horta BL, Gigante DP, Gonçalves H, et al. Cohort Profile Update: The 1982 Pelotas (Brazil) Birth Cohort Study. Int J Epidemiol 2015;44:441–441e. doi:10.1093/IJE/DYV017

22 Victora CG, Barros FC. Cohort profile: the 1982 Pelotas (Brazil) birth cohort study. Int J Epidemiol 2006;35:237–42. doi:10.1093/IJE/DYI290

23 Victora CG, Hallal PC, Araújo CLP, et al. Cohort profile: the 1993 Pelotas (Brazil) birth cohort study. Int J Epidemiol 2008;37:704–9. doi:10.1093/IJE/DYM177

24 Gonçalves H, Wehrmeister FC, Assunção MCF, et al. Cohort Profile Update: The 1993 Pelotas (Brazil) Birth Cohort follow-up at 22 years. Int J Epidemiol 2018;47:1389–1390E. doi:10.1093/IJE/DYX249

25 Bielemann RM, LaCroix AZ, Bertoldi AD, et al. Objectively Measured Physical Activity Reduces the Risk of Mortality among Brazilian Older Adults. J Am Geriatr Soc 2020;68:137–46. doi:10.1111/JGS.16180

26 Hallal PC, Bertoldi AD, Domingues MR, et al. Cohort Profile: The 2015 Pelotas (Brazil) Birth Cohort Study. Int J Epidemiol 2018;47:1048–1048H. doi:10.1093/IJE/DYX219

27 Ricardo LIC, Da Silva ICM, Martins RC, et al. Protocol for Objective Measurement of Infants’ Physical Activity using Accelerometry. Med Sci Sports Exerc 2018;50:1084–92. doi:10.1249/MSS.0000000000001512

28 da silva ICM, van hees VT, Ramires V v., et al. Physical activity levels in three Brazilian birth cohorts as assessed with raw triaxial wrist accelerometry. Int J Epidemiol 2014;43:1959–68. doi:10.1093/IJE/DYU203

29 Rowlands A v. Moving Forward With Accelerometer-Assessed Physical Activity: Two Strategies to Ensure Meaningful, Interpretable, and Comparable Measures. Pediatr Exerc Sci 2018;30:450–6. doi:10.1123/PES.2018-0201

30 Rowlands A v., Yates T, Davies M, et al. Raw Accelerometer Data Analysis with GGIR R-package: Does Accelerometer Brand Matter? Med Sci Sports Exerc 2016;48:1935–41. doi:10.1249/MSS.0000000000000978

31 Migueles JH, Rowlands A v., Huber F, et al. GGIR: A Research Community–Driven Open Source R Package for Generating Physical Activity and Sleep Outcomes From Multi-Day Raw Accelerometer Data. J Meas Phys Behav 2019;2:188–96. doi:10.1123/JMPB.2018-0063

32 van Hees VT, Fang Z, Langford J, et al. Autocalibration of accelerometer data for free-living physical activity assessment using local gravity and temperature: an evaluation on four continents. J Appl Physiol (1985) 2014;117:738–44. doi:10.1152/JAPPLPHYSIOL.00421.2014

33 van Hees VT, Gorzelniak L, Dean León EC, et al. Separating Movement and Gravity Components in an Acceleration Signal and Implications for the Assessment of Human Daily Physical Activity. PLoS One 2013;8:e61691. doi:10.1371/JOURNAL.PONE.0061691

34 Ricardo LIC, Wendt A, Galliano LM, et al. Number of days required to estimate physical activity constructs objectively measured in different age groups: Findings from three Brazilian (Pelotas) population-based birth cohorts. PLoS One 2020;15. doi:10.1371/JOURNAL.PONE.0216017

35 Hildebrand M, van Hees VT, Hansen BH, et al. Age group comparability of raw accelerometer output from wrist- and hip-worn monitors. Med Sci Sports Exerc 2014;46:1816–24. doi:10.1249/MSS.0000000000000289

36 Hill Collins P. Black feminist thought : knowledge, consciousness, and the politics of empowerment.; :357.

37 Bose CE. PATRICIA HILL COLLINS SYMPOSIUM Intersectionality and Global Gender Inequality. Published Online First: 2012. doi:10.1177/0891243211426722

38 Filmer D, Pritchett LH. Estimating wealth effects without expenditure data--or tears: an application to educational enrollments in states of India. Demography 2001;38:115–32. doi:10.1353/DEM.2001.0003

39 Hosseinpoor AR, Bergen N, Barros AJD, et al. Monitoring subnational regional inequalities in health: measurement approaches and challenges. Int J Equity Health 2016;15. doi:10.1186/S12939-016-0307-Y

40 Rowlands A, Davies M, Dempsey P, et al. Wrist-worn accelerometers: recommending ~1.0 mg as the minimum clinically important difference (MCID) in daily average acceleration for inactive adults. Br J Sports Med 2021;55:814–5. doi:10.1136/BJSPORTS-2020-102293

41 Ricardo LIC, da Silva ICM, de Andrade Leão OA, et al. Objectively measured physical activity in one-year-old children from a Brazilian cohort: Levels, patterns and determinants. International Journal of Behavioral Nutrition and Physical Activity 2019;16. doi:10.1186/s12966-019-0895-1

42 Telford RM, Telford RD, Olive LS, et al. Why Are Girls Less Physically Active than Boys? Findings from the LOOK Longitudinal Study. PLoS One 2016;11:e0150041. doi:10.1371/journal.pone.0150041

43 Telama R, Yang X, Leskinen E, et al. Tracking of Physical Activity from Early Childhood through Youth into Adulthood. Med Sci Sports Exerc 2014;46:955–62. doi:10.1249/MSS.0000000000000181

44 The Lancet Public Health. Time to tackle the physical activity gender gap. Lancet Public Health 2019;4:e360. doi:10.1016/S2468-2667(19)30135-5

45 Duffey K, Barbosa A, Whiting S, et al. Barriers and Facilitators of Physical Activity Participation in Adolescent Girls: A Systematic Review of Systematic Reviews. Front Public Health 2021;9:1529. doi:10.3389/FPUBH.2021.743935/BIBTEX

46 Chen X, Kemperman A, Timmermans H. Socio-demographics, neighborhood characteristics, time use, and leisure-time physical activity engagement patterns over the life course. SSM Popul Health 2022;19:101244. doi:10.1016/J.SSMPH.2022.101244

47 Evenson KR, Aytur SA, Borodulin K. Physical Activity Beliefs, Barriers, and Enablers among Postpartum Women. J Womens Health 2009;18:1925. doi:10.1089/JWH.2008.1309

48 Kari JT, Pehkonen J, Hirvensalo M, et al. Income and Physical Activity among Adults: Evidence from Self-Reported and Pedometer-Based Physical Activity Measurements. PLoS One 2015;10. doi:10.1371/JOURNAL.PONE.0135651

49 Beenackers MA, Kamphuis CBM, Giskes K, et al. Socioeconomic inequalities in occupational, leisure-time, and transport related physical activity among European adults: A systematic review. International Journal of Behavioral Nutrition and Physical Activity 2012 9:1 2012;9:1–23. doi:10.1186/1479-5868-9-116

50 Silva ICM da, Mielke GI, Bertoldi AD, et al. Overall and Leisure-Time Physical Activity Among Brazilian Adults: National Survey Based on the Global Physical Activity Questionnaire. J Phys Act Health 2018;15:212–8. doi:10.1123/JPAH.2017-0262

