## Supplementary material for "Gender gap for accelerometry-based physical activity across different age groups in five Brazilian cohort studies": Suppelementary material

**Supplementary table 1.** Comparison between mean overall PA and MVPA and gender for each cohort's follow-up

| Cohorts' follow-ups | Year | Overall PA (m*g*) | | | MVPA (minutes) | | |
|---|---|---|---|---|---|---|---|
| | | Men | Women | | Men | Women | |
| | | Mean (95%CI) | Mean (95%CI) | p-value* | Mean (95%CI) | Mean (95%CI) | p-value* |
| **2015 Cohort** | | | | | | | |
| 1 year | 2016 | 27.0 (26.3; 27.3) | 25.8 (25.4; 26.1) | <0,001 | - | - | - |
| 2 years | 2017 | 38.4 (37.9; 38.9) | 36.0 (35.5; 36.5) | <0,001 | - | - | - |
| 4 years | 2019 | 50.4 (49.7; 51.0) | 46.1 (45.6; 46.6) | <0,001 | - | - | - |
| **2004 Cohort** | | | | | | | |
| 6 years | 2010 | 63.8 (62.9; 64.7) | 54.7 (54.0; 55.5) | <0,001 | 74.0 (71.7; 76.3) | 50.4 (48.6; 52.1) | <0,001 |
| 11 years | 2015 | 69.9 (69.0; 70.8) | 60.1 (59.4; 60.7) | <0,001 | 55.3 (53.4; 57.1) | 30.6 (29.4; 31.7) | <0,001 |
| 15 years | 2019 | 34.7 (33.8; 35.6) | 30.7 (30.1; 31.3) | <0,001 | 39.7 (37.2; 42.2) | 26.0 (24.2; 27.7) | <0,001 |
| **1993 Cohort** | | | | | | | |
| 18 years | 2011 | 43.8 (43.1; 44.5) | 35.3 (34.9; 35.7) | <0,001 | 71.5 (69.1; 74.0) | 38.7 (37.2; 40.1) | <0,001 |
| 22 years | 2015 | 37.7 (37.0; 38.4) | 32.0 (31.6; 32.5) | <0,001 | 42.5 (40.2; 44.8) | 24.5 (23.3; 25.6) | <0,001 |
| **1982 Cohort** | | | | | | | |
| 30 years | 2012 | 38.2 (37.6; 38.9) | 33.3 (32.8; 33.8) | <0,001 | 43.6 (41.4; 45.9) | 28.5 (27.1; 29.9) | <0,001 |
| ***COMO VAI?* study** | | | | | | | |
| ≥ 60 years | 2014 | 22.0 (21.1; 22.9) | 21.5 (20.9; 22.1) | 0.328 | 13.9 (11.9; 16.0) | 7.0 (6.0; 7.9) | <0,001 |

*P-values obtained through two-tailed t-tests

**Supplementary table 2.** Intersectionality between gender and wealth: mean overall PA and MVPA gender comparison stratified by highest and lowest wealth quintiles for each cohort's follow-up

| Cohorts' follow-ups | Poorest (Q1) | | | Wealthiest (Q5) | | |
|---|---|---|---|---|---|---|
| | Men | Women | | Men | Women | |
| | Mean (95%CI) | Mean (95%CI) | p-value* | Mean (95%CI) | Mean (95%CI) | p-value* |
| ***Overall PA(mg):*** | | | | | | |
| **2015 Cohort** | | | | | | |
| 1 year | 26.3 (25.7; 27.0) | 25.9 (25.0; 26.7) | 0.359 | 27.0 (26.2; 27.7) | 25.1 (24.4; 25.8) | 0.004 |
| 2 years | 39.7 (38.4; 40.9) | 35.4 (34.3; 36.6) | <0.001 | 37.7 (36.6; 38.8) | 35.3 (34.1; 36.4) | <0.001 |
| 4 years | 52.5 (51.1; 53.9) | 47.9 (46.5; 49.3) | <0.001 | 50.4 (49.0; 51.8) | 45.2 (44.0; 46.3) | <0.001 |
| **2004 Cohort** | | | | | | |
| 6 years | 68.5 (66.1; 70.9) | 58.1 (56.0; 60.2) | <0.001 | 59.5 (57.4; 61.5) | 53.7 (52.0; 55.4) | <0.001 |
| 11 years | 76.7 (74.4; 79.0) | 64.8 (63.0; 66.6) | <0.001 | 64.9 (62.9; 66.9) | 58.1 (56.6; 59.6) | <0.001 |
| 15 years | 37.3 (35.1; 39.4) | 32.6 (30.9; 34.3) | <0.001 | 32.5 (30.4; 34.5) | 28.6 (27.4; 29.8) | <0.001 |
| **1993 Cohort** | | | | | | |
| 18 years | 47.2 (45.6; 48.8) | 37.3 (36.2; 38.4) | <0.001 | 37.5 (36.2; 38.8) | 32.0 (31.1; 32.9) | <0.001 |
| 22 years | 41.1 (39.3; 42.8) | 33.8 (32.7; 34.8) | <0.001 | 32.7 (31.3; 34.0) | 29.9 (29.0; 30.9) | <0.001 |
| **1982 Cohort** | | | | | | |
| 30 years | 40.8 (39.4; 42.2) | 36.0 (34.9; 37.2) | <0.001 | 35.9 (34.1; 37.8) | 31.6 (30.4; 32.7) | <0.001 |
| ***COMO VAI?* study** | | | | | | |
| ≥ 60 years | 22.6 (20.4; 24.9) | 20.3 (18.9; 21.6) | 0.07 | 22.7 (20.8; 24.6) | 22.0 (20.4; 23.6) | <0.001 |
| ***MVPA (minutes):*** | | | | | | |
| **2015 Cohort** | | | | | | |
| 1 year | - | - | - | - | - | - |
| 2 years | - | - | - | - | - | - |
| 4 years | - | - | - | - | - | - |
| **2004 Cohort** | | | | | | |
| 6 years | 82.1 (76.1; 88.1) | 58.2 (53.3; 63.0) | <0.001 | 66.9 (62.1; 71.7) | 45.0 (40.8; 49.3) | <0.001 |
| 11 years | 68.3 (63.3; 73.4) | 40.0 (36.8; 43.2) | 0.001 | 45.3 (41.5; 49.1) | 25.2 (22.8; 27.7) | <0.001 |
| 15 years | 52.1 (44.9; 59.4) | 36.6 (31.4; 41.7) | 0.001 | 31.7 (26.5; 36.9) | 17.0 (14.0; 20.0) | <0.001 |
| **1993 Cohort** | | | | | | |
| 18 years | 85.9 (79.9; 92.0) | 45.7 (41.8; 49.7) | <0.001 | 49.3 (45.1; 53.4) | 28.6 (26.0; 31.1) | <0.001 |
| 22 years | 54.4 (48.0; 60.8) | 29.8 (26.8; 32.9) | 0.001 | 28.5 (24.6; 32.5) | 20.3 (18.1; 22.4) | <0.001 |
| **1982 Cohort** | | | | | | |
| 30 years | 51.3 (46.3; 56.3) | 37.4 (33.4; 41.3) | <0.001 | 32.8 (27.9; 37.7) | 24.5 (21.2; 27.8) | 0.005 |
| ***COMO VAI?* study** | | | | | | |
| ≥ 60 years | 17.8 (12.0; 23.6) | 7.3 (5.0; 9.6) | 0.590 | 13.2 (9.2; 17.2) | 6.7 (4.6; 8.7) | 0.002 |

Wealth quintiles at birth for the 2015 and 2004 cohorts, at 2 years for the 1982 cohort, at 11 years for the 1993 cohort and at baseline for the COMO VAI? study

*p-values obtained through two-tailed t-tests

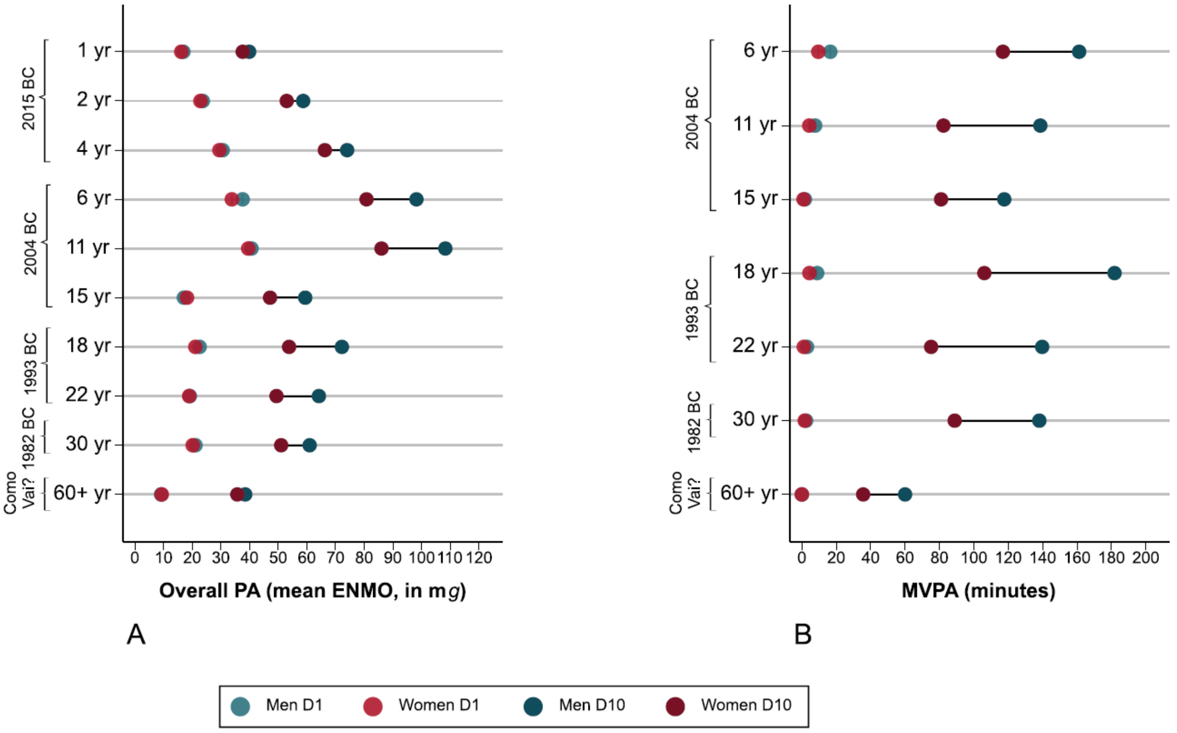


**Supplementary figure 1.** Gender gap for overall PA expressed in m*g* (A) and minutes spent in MVPA (B) across the first and last PA deciles among participants of five Pelotas cohort studies
